# Measuring integration and influence among health professionals. A social network approach

**DOI:** 10.1101/2020.04.07.20057398

**Authors:** Nelson Aguirre, Peter Carswell, Tim Keneally

## Abstract

Programs to manage long term conditions in developed countries seek greater integration and communication between health professionals to improve health outcomes. But more empirical evidence is needed to measure integration and influence for health those who are involved in these programs. This paper analyses the way in which health professionals interact at the primary and secondary care interface, based on integrated care design in a program of chronic care in New Zealand. The frequency, quality and perceived value of the interactions between general practitioners, practices nurses, medical specialists and specialist nurses are used as explanatory variables in graphical models which take a social network analysis approach. The paper shows the configuration of the network between health professionals in an integrated care context and explains the academic and policy implication of the findings.

## Introduction

Efforts to improve communication, interactions and cooperation between individuals and organizations, lead to better quality outcomes in a business context (Balkundi & Harrison, 2006; Choi, Kim, & Lee, 2010; Kilduff & Tsai, 2003; Krackhardt & Kilduff, 2008). In the healthcare context, integrated care is one such strategy (Berry & Mirabito, 2010; Ehrlich, Kendall, Muenchberger, & Armstrong, 2009; Johanson, 2010; Koshiyama, Ogawa, Tanaka, & Tanaka, 2010). Most frequently, where this kind of strategy has been applied in developed countries such as New Zealand, integrated care has been applied to managing long term conditions; in particular, care for people with diabetes (Letford & Ashton, 2010; Rea et al., 2010; Reid, 2007; Smith, 2009). Integrated care for diabetes aims to improve timely detection, treatment and metabolic control for patients with diabetes. It is grounded in the assumption that good communication and collaboration between general practitioners, nurses and specialists leads to better patient care and outcomes within and across the primary and secondary care interface (Browne, Kingston, Grdisa, & Markle-Reid, *2007*; Foy et al., 2010; McCormick & Boyd, 1994). Therefore, measuring how well integrated is an inter-professional network and describing influence within members of the network merge as indicators of good communication and collaboration among health professionals (Ahgren & Axelsson, 2005; Fattorea, Frosini, Salvatorec, & Tozzid, 2009).

A systematic review on how measure integration within integrated care had been proposed a set of criteria for measurement methods to guide future research and, also claims for practical and relatively easy methods for non-scientific users (Strandberg-Larsen & Krasnik, 2009). This paper proposes the social network approach to measure the level of integration and influence within inter-networks at the primary and secondary care interface. This perspective is important for both researchers and practitioners, but for managers responsible for implementation could become a tool for monitoring and maintenance of integrated care. The basic theoretical elements of Social network Theory will be outlined, also described previous experience implementing this approach in the health sector, and a practical application of this approach in the context of the primary and secondary care interface is described and analysed.

## BACKGROUND

### Social Network Theory for this study

The beginnings of Social Network Theory (SNT) come from the late 1930s when Moreno formulated socio-metric measurements and developed studies in dynamics of social groups (Moreno, 1934) and from Jennings’s works on leadership and influence (Jennings, 1937). A decade later, Bavelas used mathematical algorithms to depict patterns of interaction and communication within social groups (Bavelas, 1948). These studies are the basis for tenets put forward by Granovetter in 1973 on the strength of ties, and his reaffirmation in 1983, when he concludes that “the most pressing need for further development of network ideas is a move away from static analyses that observe a system at one point in time and to pursue instead systematic accounts of how such systems develop and change” (Granovetter, 1973, 1983).

Social networks are the result of interactions between “actors”, which can be individuals, teams, organizations or units of a company. The interactions between them are represented by elements such as communication, influence and affection or trust. Studies analysing social networks can describe how the actors are interrelated and can also be used to understand how these patterns of interaction could generate different patterns of performance. Studies in this area tend to be observational, and typically collect data through questionnaires or interviews. Network analysis can present graphically the ties between actors and calculate their characteristics using commercially-available software. (Adamic & Adar, 2005; Bonacich, 2008; Borgatti, Mehra, Brass, & Labianca, 2009; Carrington, Scott, & Wasserman, 2005; O’Malley & Marsden, 2008). Usually, the description of the network characteristics comes from mathematical or statistical analysis, and the relational linkages between entities are based on graphic techniques. To create a social network graph, individuals are represented as *nodes* in the network and the relationships that connect them are represented as *edges* that connect the nodes. Each edge indicates an information link between two entities. The relationship between actors is very important in SNT, because these patterns of interaction describe patterns of communication that shape patterns of collaboration, knowledge transfer and influence (Borgatti & Everett, 2006; Freeman, Roeder, & Mulholland, 1980).

### Empirical evidence of social network in organizational context

Studies on organizational social network have increased in recent years in response to interest in how the structure of interactions between actors affects outcomes. Two studies marked the beginnings of the social network approach in the organizational arena: a Harvard business school group in 1920s that used Sociograms to diagram the structure of the social interactions in the Westerns electric Company of Chicago (the famous Hawthorne study) and Kapfeerf’s work in 1972 that predicted strike activity by the workers in a factory by social network data (Kilduff & Tsai, 2003).

The scientific exploration of this topic dates back to the interest of psychologists such as Elton Mayo, who conducted several laboratory experiments in order to find the relationship between different working conditions and group productivity. But in early 1970s the analysis focused on communication patterns and performance using sociometry and representation graphic to find relations between players in a team (Cummings & Cross, 2003).

In the last two decades there are an increase number of empirical studies that apply social network approach in different sectors and organizations, such as biotechnology industries (Gordon, Kogut, & Shan, 1997; Kim & Higgins, 2007; Shan, Walker, & Kogut, 1994), chemical industries (Singh, 2005), financial companies (Granovetter, 2005; Jensen, 2007; Zaheer & Bell, 2005), and automobile, telecommunications and commercial aircraft (Carrington et al., 2005; Nohria & Garcia-Pont, 1991) between others.

The findings in organizational setting have confirmed that high-performing teams and leaders are located centrally between groups within the network. Among the most representative findings, direct and indirect links provide access to people who provide support and resources within their networks.

### Empirical evidence of social network in healthcare context

Recent empirical research in the healthcare context have used a social network approach to study interactions between health professionals, describing and establishing comparisons between networks using networks metrics to explore the structure and dynamic of the networks, identifying the effects of demographic and cognitive diversity on relationships, analysing the effects of homophily on network development, and assessing influence and power within the networks (Burt, Kilduff, & Tasselli, 2013; Fattore & Salvatore, 2010; Fattorea et al., 2009; Keating, Ayanian, Cleary, & Marsden, 2007; Lewis, Baeza, & Alexander, 2008; Mascia, Cicchetti, Fantini, Damiani, & Ricciardi, 2011; McDonald, Jayasuriya, & Harris, 2011; Meltzer et al., 2010; Quinlan & Robertson, 2010; Scott et al., 2005).

Researchers have also used this approach to learning about patterns of communication and information exchange within the individuals, units and organizations in emergency departments (Benham-Hutchins & Effken, 2010; Houghton et al., 2006; Patterson et al., 2013), within primary and secondary interface modelling different scenarios of patient demand to establish effects in efficiency and accessibility of the system (Farinha, Oliveira, & Sa’, 2008), and in hospitals, in order to compare the effects of the interaction between inter-professional teams across the units, to better understand collaboration within hospitals, to examine advice-seeking, diffusion of innovation and knowledge sharing between health professionals (C. Anderson & Talsma, 2011; J. G. Anderson & Jay, 1985; Boyer, Belzeaux, Maurel, Baumstarck-Barrau, & Samuelian, 2010; Creswick & Westbrook, 2010; Jippes et al., 2010; Mascia, Vincenzo, & Cicchetti, 2012; Wiemken, Ramirez, Polgreen, Peyrani, & Carrico, 2012).

Social Network theory can be applied to the social sciences as the basis for understanding the structure, cause and consequence of social interactions among members involved in the network (Scott & Carrington, 2011). Social network analysis (SNA) comprises a group of methods used for mapping, measuring and analysing the social relations between individuals, groups or organizations (Carrington et al., 2005). SNA allows the analysis of patterns of interaction among network members, that facilitates or constrains the decisions and actions of members (Blanchet & James, 2011). Previous research pointed out that social network approach provides valuable ways to assess social structures within inter-professional networks in healthcare; one of the advantages is to measure integration within the network. Integrated care is based on collaboration, communication and coordination principles. Therefore, measuring integration through social network approach provides potential benefits for health professionals, healthcare managers, policymakers and researchers.

## RESEARCH DESIGN AND METHODS

### Research Context

Counties Manukau is an area of over 350,000 residents covered by a tax-based system providing universal access to medical services in Auckland New Zealand. Secondary care services are free to the user, and primary care is subsidised from general taxation. Organizations such as Counties Manukau District Health Board (CMDHB) and Middlemore Hospital receive funding from the Ministry of Health. CMDHB contracts Primary Health Organizations (PHO) who reimburses general practices and physicians for their services (Gribben, 2003).

Counties Manukau was faced with an increased demand in secondary care services in the late 1990s, due to increased health needs of the population and particularly as a result of poor service coordination between the primary and secondary care interface. One of the critical factors of demand is concentrated in the high prevalence of chronic diseases such as diabetes and its complications. According to recent statistics, an estimated 8% of the population in this area has diabetes. The prevalence is concentrated in 15% Pacific people, 10% Maori, 8% Asian groups and 5% in those of European and other ethnic groups. Counties Manukau accounts for 14% of people with diabetes across NZ, which is why it ranks as an important area of action nationally (New Zealand office of the Auditor-General, 2007).

Since 2001, improving health system performance has been identified as a critical success factor in NZ, including CMDHB. Developing and implementing a strategic plan for the best use of human and material resources to improve coordination and integration of health care providers is a priority. Chronic care management (CCM) is the result of a set of political and organizational efforts that have been developed to address the challenges of the increasing demand of secondary care.

The problem of integrating secondary care (hospital based) with primary care providers is a central concern in integrated care. As noted, diabetes is a disease of high prevalence in this geographical area and integrated care has been configured as a strategy to respond to the challenge. The aim of this research is to explore how integrated networks of professionals at the primary and secondary care interface have developed.

### Design

Data for a social network analysis were collected using a cross-sectional survey conducted in March 2012.

### Participants

We invited people from primary care and from secondary care who were involved in the care of the same group of diabetic patients. The first group consisted of general practitioners (GP) and practice nurses (PN). The second group included endocrinologists and diabetes nurse’s specialist (DSN) who work within the secondary care diabetes service. For the first group we invited all GPs who were in Counties Manukau and members of the PHO and the second group was contacted directly at the main referral hospital at the same area.

### Procedures

This study received ethics approval from The University of Auckland Human Participants Ethics Committee and the New Zealand Ministry of Health’s Northern Regional Health and Disability Ethics Committee NTX/11/EXP/150 dated 19/07/2011. Participants completed a social network questionnaire delivered online. Reponses were collected electronically. Data were checked, cleaned, and then analysed in dedicated software (SPSS V19). The information collected included demographic characteristics, the role relationship between the respondent and each of his or her colleagues – known as “alters” in the network literature– and the frequency, quality and perceived value of the interactions between health professionals.

### Analysis and tools used

Social Network Analysis requires to define the level of analysis. Therefore, interaction within health professionals taking medical decisions in patients with diabetes were used in order to identify the level of integration and influence at the primary and secondary care interface. Interactions are justified by the need for communication, coordination and collaboration, based on the recognition that there are differences on the influence and power that each actor posed in the network (some actors are “seekers” while others are “providers”). Therefore, identifying important actors in a network allow understanding influence within a network (Wasserman & Galaskiewicz, 1994). However, health professionals take part simultaneously in different networks for care patient with diabetes at this interface. For example: health professionals share interactions within the general practice, across general practices or within multidisciplinary teams. Including or excluding interactions of one or more of these functional networks may change the results of analysis of a specific social network. As a result, I have defined two units of analysis with the following boundaries: A first network (network A) that represents the whole interface: it is represented by interactions and sub-networks in both sides of the interface (primary care: within and across general practices and secondary care: within health professionals) and also interactions across the sectors. A second network (Network B) that only represents interactions between primary and secondary care (across the sector) and excluding interactions within secondary care. Excluding the effect of ties between endocrinologist and specialist nurses prevents the effect of the high density of interactions between these two health professional groups from obscuring the less dense network between primary and secondary care.

Social Network measurements (table 1) are the way to describe social interactions that comprise the network. These are indicators which reflect the differences and similarities of interactions between actors according to their position within the network. (Carrington et al., 2005; Marsden, 1990). All of these indicators and network maps were calculated and performed using UCINET V6 and GEPHY V0.8.2-Beta

**Table 1.** Summary of Social Network Measurements for this study

|  | Measure | Definition | Meaning | Usage |
| --- | --- | --- | --- | --- |
| **Network Level Measures** | *Density* | The proportion of observed relationships amongst all possible relationships | Speed of information flow | Identifying the general level of connectivity and integration across the network |
|  | *Geodesic distance* | The short distance (minimum number of paths) separating a pair of actors | Speed of information | Speed of diffusion of ideas; |
| **Centrality Measures** | *Degree centrality* | Node with the most connections | In the know | Identifying sources; reducing information flow |
|  | *Betweenness* | Node in the best paths | Connects groups | Typically has political influence, but may be too constrained to act |
|  | *Eigenvector centrality* | Node most connected to other highly connected nodes | Strong social capital | Identifying those who can mobilize others |
|  | *Closeness* | Node that is closest to all other nodes | Rapid access to information | Identifying sources to acquire/transmit information |
|  | *Betweenness – centrality* | High in betweenness but not degree centrality. | Connects disconnected groups | Go-between; reduction in activity by disconnected groups |

**Table 1:**
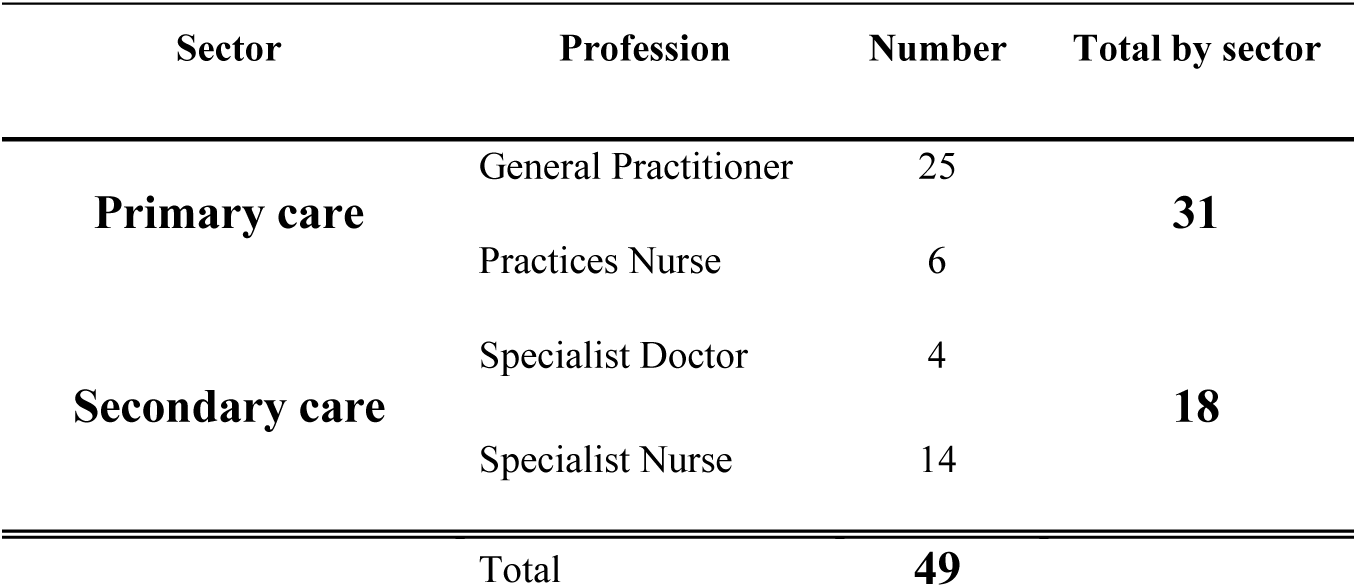
Participant’s distribution by profession and sector.

We focused the social network analysis on “density” measurement. It is a common measure to show how well connected a network is, in other words how well integrated a network is. Mathematically, it is the ratio of the number of edges in the network over the total number of possible edges between all pairs of people. We have used this measurement to evaluate the quantity of interactions that arise when making each one of the questions of the questionnaire, through this way we identified the density of interactions in three different dimensions: frequency (daily, weekly, monthly or less than monthly), quality (Interaction with a professional provides current and valuable information) and perceived value of interaction (interaction with a professional influences decisions about how diabetic patients are diagnosed and treated), As a difference of the previous analysis, the value of the interactions is measured in terms of how interactions with other health professional influence the decision about how diabetic patients are diagnosed and treated. That means high perceived value implies strong influence on the way that health workers diagnosed and treated patient with diabetes.

## RESULTS

Fifty people responded to the social network survey; one was incomplete leaving 49 who contributed to further analyses. The study had 49 participants in the social network survey from 50 invitations submitted, with a gender distribution of 65% female and 35% male. The distribution of the participants by profession and sector is shown in table 2.

**Table 2:** Social Network Measures. Network B interface

| Id | Degree | In degree | Out Degree | Closeness Centrality | Betweenness Centrality | Eigenvector Centrality |
| --- | --- | --- | --- | --- | --- | --- |
| SCS003 | 18 | 5 | 13 | 1.5417 | 115.9286 | 0.579136 |
| SCS009 | 15 | 4 | 11 | 1.7917 | 143.7333 | 0.933615 |
| SCN010 | 15 | 4 | 11 | 1.7083 | 81.92857 | 0.647533 |
| GD00628 | 12 | 5 | 7 | 1.8333 | 131.0024 | 0.982938 |
| GD00332 | 9 | 7 | 2 | 2.5833 | 73.71667 | 1 |
| GD00514 | 9 | 4 | 5 | 2.1667 | 49.14524 | 0.521964 |
| GD00310 | 7 | 4 | 3 | 2.4167 | 44.78571 | 0.714413 |
| SCN001 | 7 | 1 | 6 | 2.125 | 26.23333 | 0.314896 |
| SCN011 | 7 | 2 | 5 | 3.125 | 19.66667 | 0.563052 |
| GD00926 | 6 | 4 | 2 | 2.6667 | 22.35 | 0.893294 |
| GD00335 | 6 | 0 | 6 | 2.1481 | 0 | 0 |
| GD00832 | 6 | 6 | 0 | 0 | 0 | 0.996717 |
| GD00629 | 5 | 3 | 2 | 2.5833 | 16.66667 | 0.71113 |
| GD00427 | 5 | 5 | 0 | 0 | 0 | 0.901136 |
| SCN013 | 4 | 1 | 3 | 2.92 | 1.642857 | 0.003283 |
| GD00122 | 4 | 4 | 0 | 0 | 0 | 0.714413 |
| GD00731 | 4 | 4 | 0 | 0 | 0 | 0.81127 |
| SCS011 | 4 | 4 | 0 | 0 | 0 | 0.784627 |
| GD00312 | 3 | 2 | 1 | 2.5 | 6.333333 | 0.207431 |
| SCN012 | 3 | 2 | 1 | 2.7917 | 3.866667 | 0.545081 |
| GD00930 | 3 | 3 | 0 | 0 | 0 | 0.71113 |
| SCN008 | 3 | 3 | 0 | 0 | 0 | 0.72896 |
| GD00114 | 2 | 2 | 0 | 0 | 0 | 0.103423 |
| GD00512 | 2 | 2 | 0 | 0 | 0 | 0.381752 |
| GD00535 | 2 | 0 | 2 | 2.4231 | 0 | 0 |
| SCC005 | 2 | 0 | 2 | 3.4 | 0 | 0 |
| SCN003 | 2 | 2 | 0 | 0 | 0 | 0.318179 |
| SCN004 | 2 | 0 | 2 | 3.16 | 0 | 0 |
| SCN007 | 2 | 0 | 2 | 3.4 | 0 | 0 |
| SCN014 | 2 | 2 | 0 | 0 | 0 | 0.006566 |
| GD00630 | 1 | 1 | 0 | 0 | 0 | 0.297148 |
| GD00723 | 1 | 0 | 1 | 2.72 | 0 | 0 |
| SCN009 | 1 | 1 | 0 | 0 | 0 | 0.17932 |

**Table 3.**
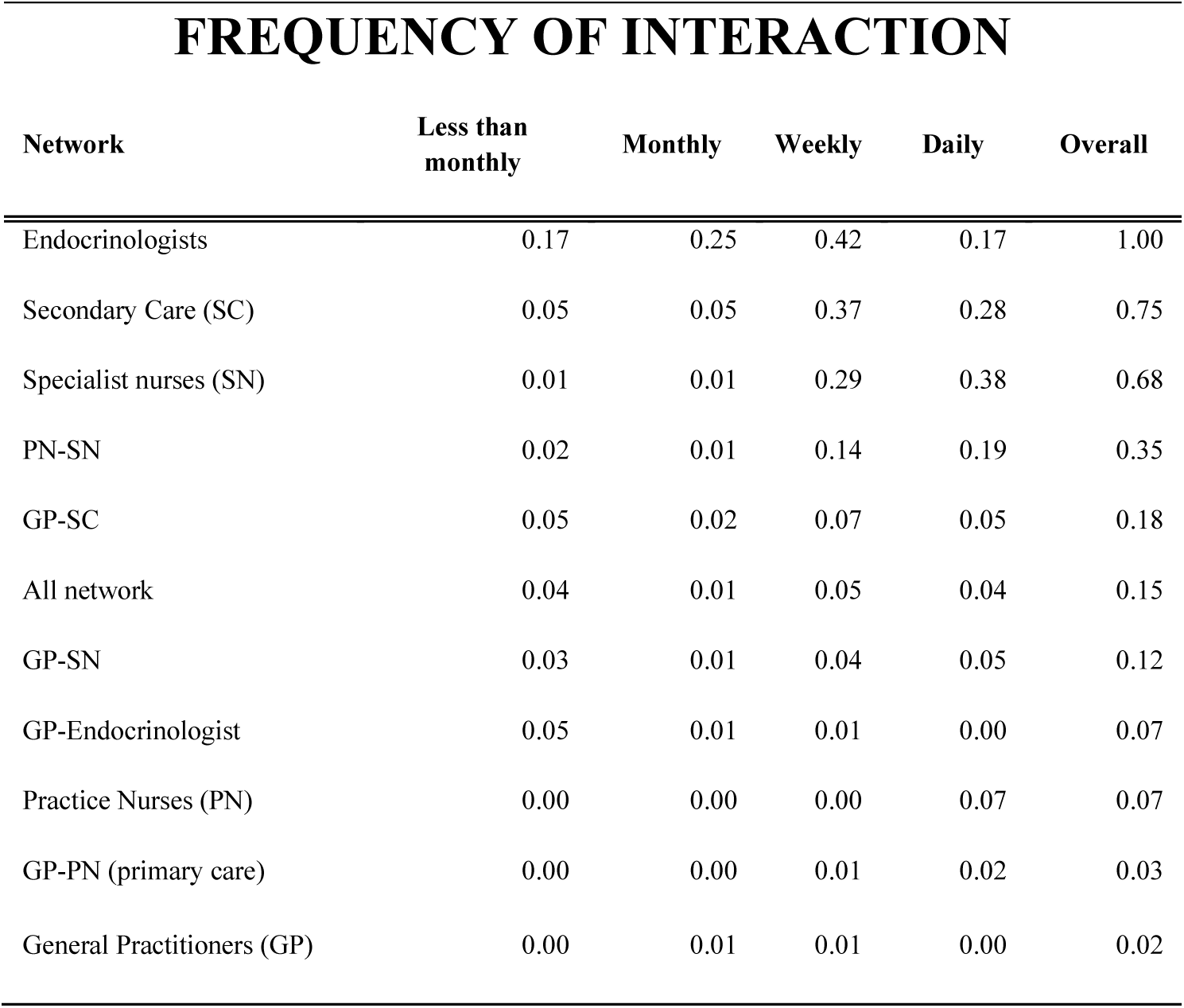
Frequency of interaction about diabetes care at the primary and secondary interface. Numbers represent the density measure in SNA when it was selected each network. Higher numbers indicate higher density of interaction within the group of health professionals at the particular frequency of interaction.

A total of 18 health practices from primary care were represented by at least one participant. Figure 1 shows network A and represents how heath workers were connected within and between primary and secondary care. The greater size of the spheres and a more central position on the network represents greater relative influence. The “star” graph suggests that health workers in positions more central of the network exert more dense interactions and influence over the rest of the inter-professional network. In the middle of the graph were a dense group of specialist nurses and endocrinologists (red spheres) interacting for the purpose of providing support and advice to general practitioners and nurses (blue spheres). Density of interactions within this network is 15.2, which is relatively low.

**Figure 1:**
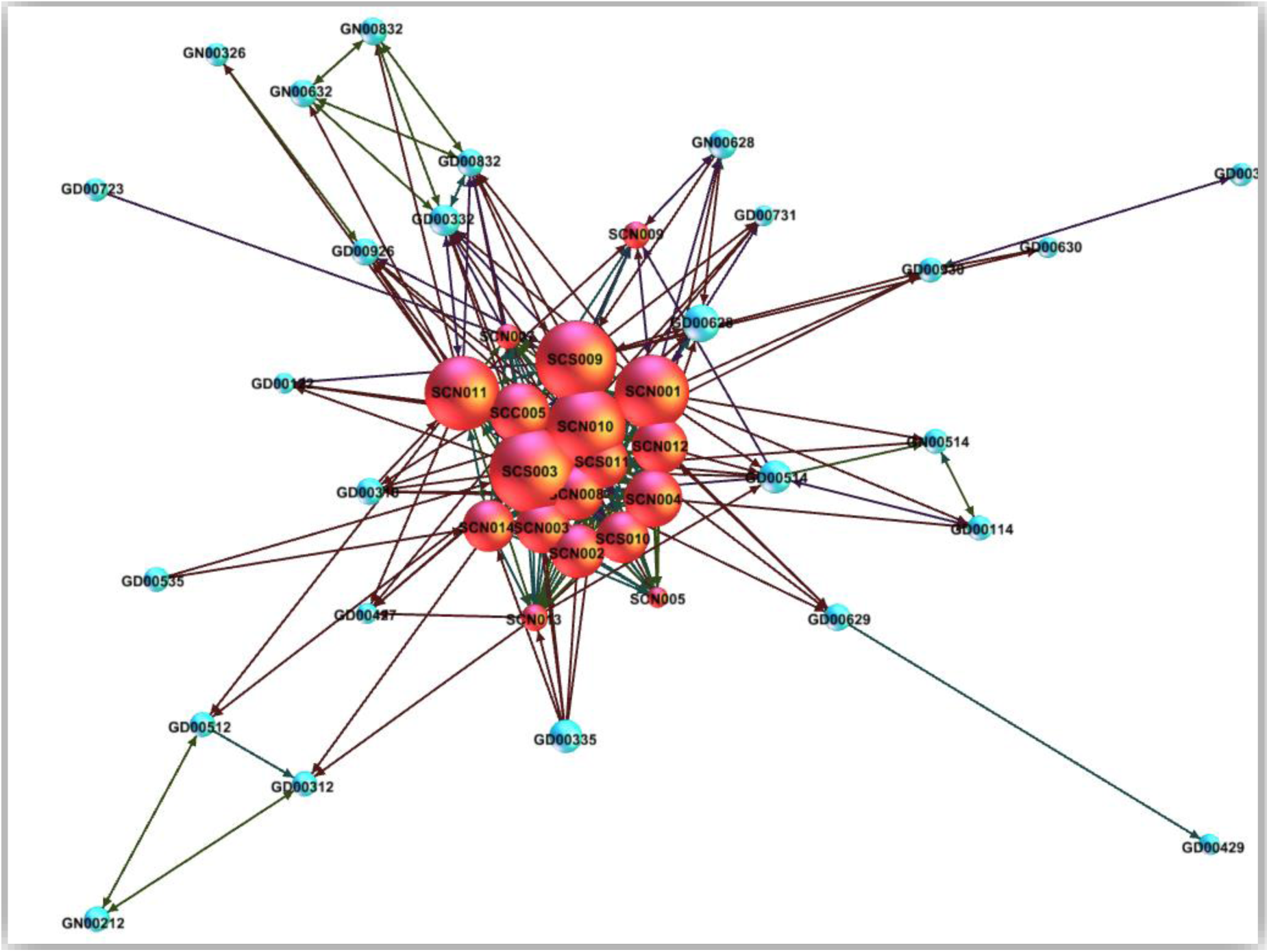
Network A, all primary care and secondary care interactions. Blue: Primary care sector & Red: Secondary care

Figure 2: network B map. Interactions between Primary and secondary care by profession and within primary care (omitting interactions within secondary care).2 shows the distribution of the network at the interface between primary and secondary care after excluding interactions solely within secondary care.

**Figure 2:**
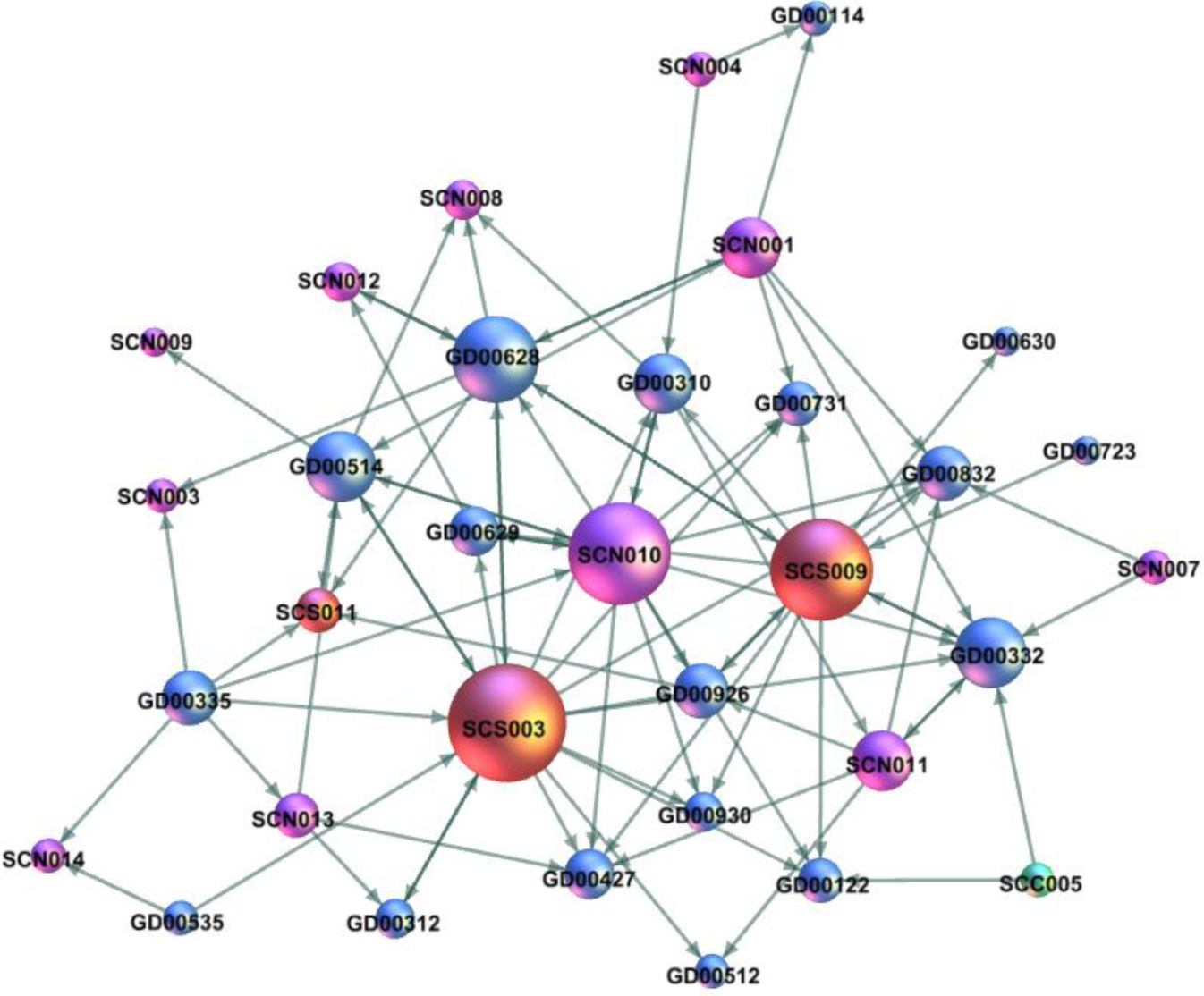
network B map. Interactions between Primary and secondary care by profession and within primary care (omitting interactions within secondary care). Red: specialist, Blue: GP and purple and Green: DSN.

This figure shows the importance and centrality of three secondary care professionals (SCS003, SCS009 and SCN010) in this network, and contrasts with network A in that DNSs are more peripheral. This graph illustrates the relatively higher centrality of some five GPs (GD00628, GD00332, GD00514, GD00326 and GD00926) who relatively were more connected with both secondary care professionals and with primary care colleagues.

The *density* of the interactions in network B has reduced to 3.7%, as the interactions between secondary care’s health professionals were eliminated. Table 2: Social Network Measures. Network B interface3 shows the social network measurements at the interface’s network. The overall results are consistent with the map shown in Figure 2: network B map. Interactions between Primary and secondary care by profession and within primary care (omitting interactions within secondary care).2. Three secondary care health professionals possess the highest *degree* and *betweenness* in both Networks A and B. Because *closeness centrality* is one measure of strength of interaction between GPs and secondary care professionals, it appears that professionals GD0047, GD0630 and GD0723 have weakest relationships with secondary care. However, high *eigenvectors* shows that two GPs (GD0047 and GD0630) are strongly connected through leaders in the network.

Comparing the two Network figures shows three important elements. First, the structure of network A is a result of the great density of interaction between health professionals in secondary care and between DNSs. Second, endocrinologist are the most central or important actors in both networks, followed by DNSs. The social network questionnaire asked if an interaction with a particular health professional influences his/her about how diabetic patients are diagnosed and treated, and also if this interaction provides current and valuable information that influences decisions about how to care patients with diabetes, which means that the network structure suggested the direction of the influence and advice across the interface. Third, there are a range of differences in centrality measures within the GPs, some of them appear well and strong connected within the network but some appear relatively isolated.

Table 4 illustrates the distribution of the network densities by frequency of interaction in relation to their profession. Each value represents the calculated density of interaction obtained when a group of actors interact in a particular frequency. In general there is a higher frequency of interaction within secondary care (0.75) than in primary care (0.03) on issues related to diabetes. Endocrinologists interact overall more about diabetes than the rest of the network followed by DNSs. Endocrinologists have more dense interaction weekly while DNSs do on a daily basis. There is a more frequent communication between nurses (PNs and DNSs), higher than communication with GPs with secondary care. As in this study frequency of interaction represents advice regarding how to treat patients with diabetes, the data reveal that GPs seeks such advice from secondary care rather than from other GPs. The frequency of interaction for general practitioners with secondary care is weekly and higher in contrast to the frequency of interaction between GPs which is weekly or monthly. Interaction between GPs and DNSs were more frequent than between GPs and endocrinologists. Physicians (GP and endocrinologist) interactions can be less than monthly, while the interactions between GPs and DNSs can be weekly or even daily.

**Table 4.**
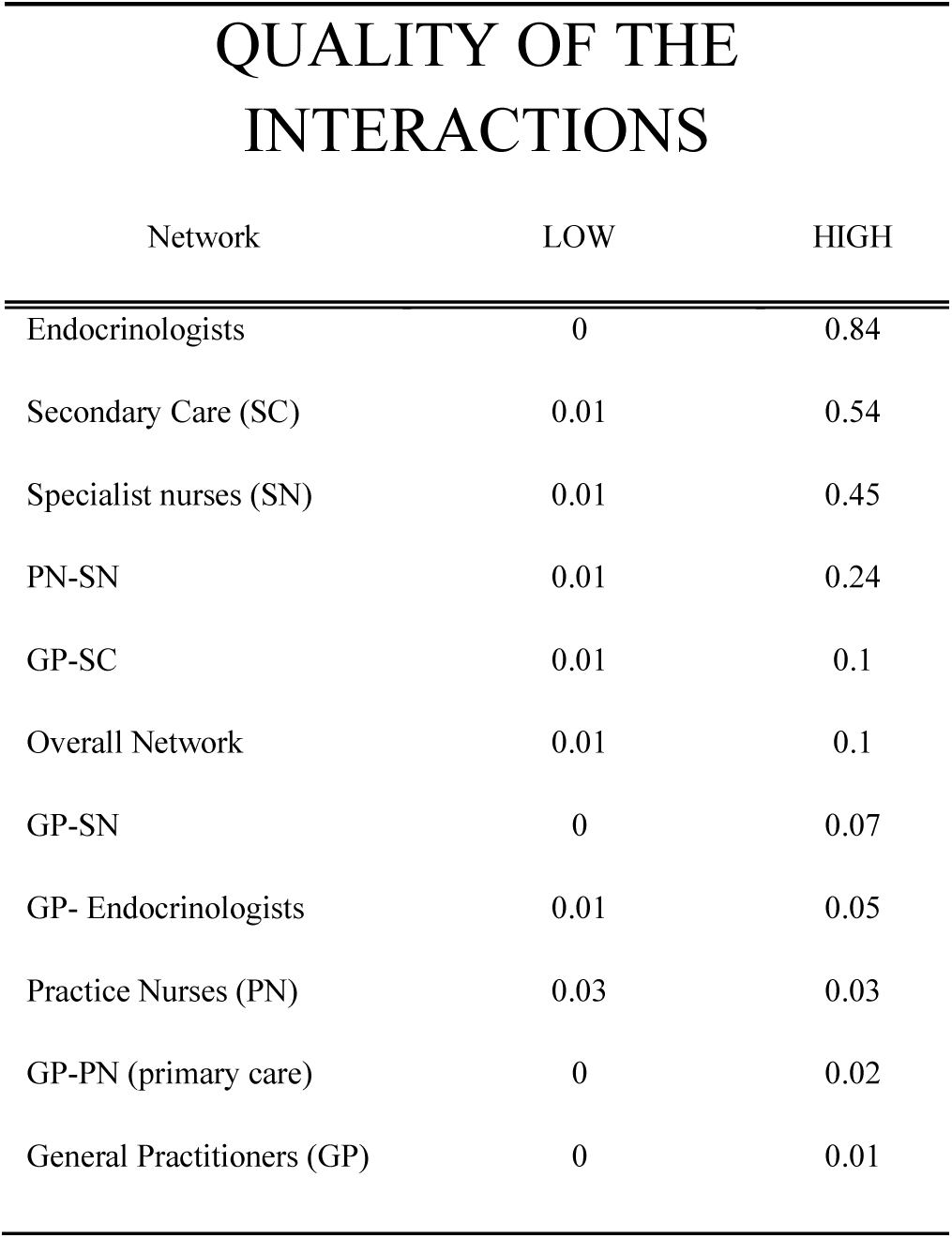
Quality of interaction about diabetes care at the primary and secondary interface

Table 5 shows that interactions between endocrinologists, followed by interactions within secondary care, were ascribed with high quality (84% and 53% respectively). In contrast, interactions within GPs were considered high quality in just 1%. Therefore, if GPs seek advice, they were more likely to value discussion with professionals in secondary care than colleagues at the same practice or elsewhere in primary care. However, the quality of the interaction between GPs and DNSs was ranked higher than interactions with endocrinologist, and 1% of the density of the interactions was ranked low quality. That means there are factors to affect the quality of the interactions between GP and endocrinologist related to current and value of the information. For DNSs and PNs ascribe higher quality to the information they share than do GPs when they interact with secondary care.

**Table 5.** Perceived value of interaction about diabetes care at the primary and secondary interface

| PERCEIVED VALUE OF THE INTERACTIONS |  |  |
| --- | --- | --- |
| Network | LOW | HIGH |
| Specialist | 0 | 0.84 |
| Secondary Care (SC) | 0.01 | 0.53 |
| Diabetes specialist nurses (DSN) | 0 | 0.44 |
| Nurses (DNSs and PN) | 0.01 | 0.23 |
| GP-SC | 0.01 | 0.11 |
| Overall Network | 0.01 | 0.1 |
| GP-SN | 0.01 | 0.06 |
| Doctors (GPs and Endocrinologist) | 0.02 | 0.05 |
| Practice Nurses (PN) | 0.03 | 0.03 |
| GP-PN (primary care) | 0.01 | 0.02 |
| General Practitioners (GP) | 0 | 0.01 |

Table 5. Perceived value of interaction about diabetes care at the primary and secondary interface6 shows the most perceived value of the interactions is hold within secondary care (endocrinologist and specialist nurses). In contrast with the low perceived value of the interactions within primary care.

Although GPs considered interactions with endocrinologist with high perceived value there is still a percentage of the interactions ranked with low perceived value. Therefore GPs considered the information from endocrinologist is current and valuable (high quality) but in some cases does not help to take decisions on how treat patient with diabetes.

## DISCUSSION

The social network perspective applied to the primary and secondary care interface is one of the contributions of this study. Defining two Networks proved a useful way to confirm that the centrality of secondary care and its influence on primary care is real rather than an artefact of the high density of interactions between endocrinologist and DNSs. The selected social network measures constitute measures of integration within the current health professional’s network and, anticipate information to analyse potential issues for knowledge transfer and advice. It has been suggested that the small size of networks in some chronic care programs could be a critical barrier to patient care because of the limited resources and capacities that each actor has for building and maintaining ties (Scott & Carrington, 2011), allowing access to information from valuable collaborators (Burt, 2009), and also the introduction of new practices and behaviours reinforcement (Valente & Fujimoto, 2010). In regard to the centrality measures, *in-degree* may be especially important as it reflects the influence and importance of individuals. Diabetes care at the interface between primary/secondary care recognizes actor’s influence based on their specialized knowledge and expertise (opinion leaders).

Some GPs in this study showed 100% density of interaction with his/her colleagues within the general practice, and also has many interactions and connections with secondary care professionals. Those GPs could share information with important players in secondary care as well as with his/her colleagues in primary care, such that otherwise-isolated health professionals could indirectly receive the benefits of interaction with secondary care. This represents a structural hole as two separate clusters (general practice and secondary care professionals) possess non-redundant information, but a mutual connection to allow flow of information. Some health professionals in primary care appear to be disconnected from the influential actors and their specialised knowledge. However, selecting the interface network showed that some of these GPs apparently isolated had interactions with other actors that were connected with influential actors. This finding is very important in terms of access to information and knowledge.

Overall analysis confirms Zappa’s (2011) findings related to the heterogeneity in the tendency to build interactions with other colleagues (Zappa, 2011). A phenomena seeing other networks and in different contexts (Freeman, 2008; Nohria & Eccles, 1992; Scott & Carrington, 2011; Wasserman & Galaskiewicz, 1994). This variability represents differences in motivation and interest for building interactions within a social network in order to solve personal needs and is associated with the decision to adopt innovations or new knowledge (Kim & Higgins, 2007; Zappa, 2011). Some actors seek advice when they encountering an issue or opportunity to resolve individual doubts, and as a result actors able to share relevant knowledge and information have more central positions in the network (Nebus, 2006).

The current results presents an opportunity to take in account two different interpretation of density measures: Coleman (1988) states that dense networks are the optimal social structure, while Burt (2009) states that disconnected alters is the optimal strategy for a network. Coleman and others would argue that higher network density implies better access to timely information for primary care (Coleman, 1988; Gulati & Gargiulo, 1999; Rowley, Behrens, & Krackhardt, 2000). This could be particularly evident for complex knowledge, as this requires direct, frequent and intense relationship. In contrast with this position, Burt (2009) considers that density may affect the diffusion of information *within* the groups in a network rather than *between* groups in a network. As a result structural holes within the network offer one effective way to share information. This explanation could apply in this context as GPs and PNs do not work alone, they share work places and patients within a general practices. Communication and information is relatively easy to maintain within practices as there are a variety of ways and opportunities of interaction: i.e. clinical meetings, lunch time, informal corridor meetings, and virtual patient clinical data on the local system are all examples provided by participants during interviews. As a consequence of these forms of interaction, professional’s thinking may be homogeneous among health professionals within health practices, as people converge in beliefs and practices developed within the group. In fact, they could attempt to block different views of information from external sources, if they consider the information is not concordant with their perspectives or interest.

However, novel and complex knowledge comes from specialised and expert health professionals at the external setting. Secondary care represents one of the main sources to obtain this kind of knowledge and, the current results have shown low density of interaction between primary care groups and secondary care. Burt would see this as an opportunity to build effective networks. Some of the health workers in general practices do not have strong relationship with specialised people. But it does not mean they are not aware of the existence of secondary care. It means people are focussed on their professional daily life activities and they do not attend the activities from other different groups. As a result there are gaps or empty spaces within the inter-professional network at this interface. GPs have to deal with patients with a great range of diseases and complications; not all GPs or PNs are likely to be equally involved or even interested specifically in diabetes care. Fortunately, other GPs or PNs more involved in diabetes care are willing to invest time and resources to build ties with expert people and then spread and share this new knowledge with their colleagues in primary care.

Social Network Analysis could be a useful tool to measure levels of integration at the integrated care programs. Managers, policy makers and staff could use this approach for the purpose of measure the structure and dynamic of the inter-professional networks, the similar way as it were described and analysed in this work. The maps and mathematical analysis showed several social network characteristics in which this network performed and interact.

This measurement model applied in this study is limited to the functional aspects of the integration within integrated care. However, it is possible to measure the synergy of the integration or integration efficacy, in other words how well-functioning is likely to influence patient outcomes by the way health professionals interact.

The present study has a number of limitations that need to be take in account when interpreting its results. First, a cross-sectional study limits the possibility to draw conclusions that imply causality. Second, the size of the sample and the particularities of the context involve generalization issues. However transferability should be considered. Third, potential limitations implicit in the social network questionnaire should be considered as other studies that used this tool.

## Conclusion

The literature suggests that the strength of the ties between people in a network facilitates knowledge transfer and the diffusion of novel information. If this is a critical issue for improving patient outcomes at the primary and secondary interface in the chronic care programs, it is also critical to identify how patterns of social network characteristics affect patient outcomes. The future of the research will focus on identifying how interactions between health professional might affect patient outcomes at this interface

Social network analysis as a method used in this study offers a graphic model and information to show how health workers are interacting across the primary and secondary care interface. The density of interactions across the interface between primary and secondary care in the context of diabetes care is relatively low compared with the level of interaction within secondary care. There are differences between how health practices interact with secondary care, who doctors interact with and who nurses interact with. This may indicate different channels that the two groups operate through in developing networks. Specialist nurses are more likely to be connected with the rest of the network, and they perceive the quality and value of these interactions as a critical factor in making decision with patients with diabetes. Recommendations arising from this empirical evidence include investigating opportunities and systems for supporting general practices to interact more frequently with secondary care in order to improve health outcomes for patients.

## Data Availability

No data is available

## Acknowledgements

The authors would like to thank members of the Procare Primary Health Care in Auckland and Counties Manukau District Health Board. We appreciate the cooperation of Dr. Brandon Orr-Walker and all members of the participating GPs, Nurses, Specialists and Diabetes Nurses Specialist in Counties Manukau.

## Notes

### Competing Interest Statement

The authors have declared no competing interest.

### Funding Statement

No external funding was received for this research

